# Penetrance of Parkinson’s disease in *GBA1* carriers is depending on the variant severity and polygenic background

**DOI:** 10.1101/2025.01.23.25320896

**Authors:** Emadeldin Hassanin, Zied Landoulsi, Sinthuja Pachchek, NCER-PD Consortium, Peter Krawitz, Carlo Maj, Rejko Krüger, Patrick May, Dheeraj Reddy Bobbili

**Author notes:** These authors contributed equally to this work and share first authorship (Emadeldin Hassanin, Zied Landoulsi, Sinthuja Pachchek). These authors contributed equally to this work and share last authorship (Rejko Krüger, Patrick May, Dheeraj Reddy Bobbili). **Corresponding author:** Dheeraj R. Bobbili, Ph.D., Luxembourg Centre for Systems Biomedicine, University Luxembourg 6 avenue du Swing, L-4367 Belvaux, Luxembourg.

## Abstract

**Background:** Heterozygous variants in the *GBA1* gene cause Parkinson’s disease (PD) with variable penetrance and have been classified into severe, mild, and PD-specific risk variants based on their association with Gaucher’s disease (GD; mild and severe) or PD (risk variants). Polygenic risk scores (PRS) further modify PD susceptibility and may influence the age of onset in *GBA1* variant carriers. Our study investigates the interaction between a genome wide PRS and pathogenic *GBA1* variants (*GBA1*_PVs_), focusing on how established combined PD risk polymorphisms may influence *GBA1*-related PD risk across different levels of *GBA1*-mediated pathogenicity.

**Methods:** *GBA1* variants were identified from whole exome sequencing data in the UK Biobank (UKB) cohort and from *GBA1*-targeted PacBio sequencing in the Luxembourg Parkinson’s Study (LuxPark). PRSs were calculated for all participants using established genome-wide significant SNPs, excluding variants within the *GBA1* locus, and then categorized based on both PRS levels and *GBA1*_PVs_ carrier status. Carriers of *GBA1*_PVs_ were further divided into ‘severe (Gaucher-related) +mild (PD-related)’ and ‘risk’ groups. To evaluate the relationship between PRS, *GBA1*_PVs_ carrier status or severity, and PD risk, logistic regression and Cox proportional hazards regression were conducted with disease presence as the dependent variable.

**Results:** We identified *GBA1*_PVs_ in 8.8% of PD patients in the UKB discovery cohort and 9.9% in the LuxPark replication cohort. *GBA1*_PVs_ carriers had consistently higher PD risk compared to non-carriers across all PRS categories. In UKB, *GBA1*_PVs_ carriers in the highest PRS category had a 2.3-fold increased risk of PD (OR: 2.34; 95% CI, 2.08-2.63) and cumulative incidence of 67% by the age of 75, while those in LuxPark had a 1.6-fold higher risk (OR: 1.64; 95% CI, 1.52-1.76), and cumulative incidence of 81% at the age of 75. Carriers of “severe+mild” *GBA1* variants had nearly double the risk of PD compared to “risk” variant carriers, with ORs ranging from 2.05 to 3.69 in UKB and 1.73 to 1.98 in LuxPark. The interaction between the PRSs and *GBA1*_PVs_ severity was similar in the two cohorts.

**Conclusions:** Our findings demonstrate that *GBA1*_PVs_ carrier status and severity significantly impact PD risk, with severe variants conferring higher risk than risk ones. Additionally, PRS consistently increases both PD risk and *GBA1*_PVs_ penetrance in an additive manner across all variant types, defining a genetic background that influences PD penetrance in *GBA1*_PVs_ carriers. The presence of additional PD-associated risk variants in *GBA1* carriers defines new avenues to incorporate PRS and genetic risk data into future clinical trial design and genetic counselling in *GBA1*-associated PD.

## Introduction

Pathogenic variants in the *GBA1* gene encoding the lysosomal enzyme glucocerebrosidase (GCase) are the most common genetic risk factors for Parkinson’s disease (PD) (Sidransky et al. 2009; Parlar et al. 2023). Initially associated with Gaucher’s disease (GD) in its biallelic form, *GBA1* variants also increase PD risk in heterozygous carriers. *GBA1* variants are prevalent in 5-20% of PD patients in different populations worldwide, with a recent study identifying a common intronic variant in 40% of African PD patients (Rizig et al. 2023). The risk of developing PD follows a gradient, with severe GD-causing variants (e.g., L483P) raising PD risk 9-to 10-fold, while mild variants (e.g., N409S) increase PD risk approximately 4-fold (Höglinger et al. 2022; Parlar et al. 2023). Additionally, non-GD-causing *GBA1* variants, such as E365K and T408M, are more frequently observed in PD patients and are considered significant risk factors for PD (Höglinger et al. 2022; Parlar et al. 2023). *GBA1*-associated PD (*GBA1*-PD) is typically characterized by earlier onset, more rapid progression, and a higher frequency of non-motor symptoms (Gan-Or et al. 2018).

The penetrance of *GBA1*-PD is variable, with few carriers developing the disease only at the age of 80 (Anheim et al. 2012), conferring to a maximal penetrance of approximately 30% (Balestrino et al. 2020) complicating risk assessment. Therefore to define individual risk, it is essential to understand the role of a ‘genetic background’ as defined by common variants associated with PD risk and identified by genome-wide association studies (GWAS) (Nalls et al. 2019) among *GBA1* carriers, such as the polygenic background has been found to modulate the PD risk of *GBA1* variants in carriers of p.E365K, p.T408M and p.N409S variants, and decreasing the age of onset (AAO) of PD (Blauwendraat et al. 2020). Moreover, variants in the PD genetic risk score were more frequent in GD patients who developed PD, suggesting that common variants may play a role in shared biological pathways underlying both conditions (Blauwendraat et al. 2023).

Our study extends previous research by analyzing the impact of polygenic risk scores (PRS) across a broader range of *GBA1* pathogenic variant carriers (*GBA1*_PVs_) in the UK Biobank (UKB), with validation in the Luxembourg Parkinson’s Study (LuxPark), focusing on how PRS and *GBA1*_PV_ severity influence PD risk.

## Methods

### The UK biobank cohort

UKB is a large, long-term prospective study comprising over 500,000 participants (Bycroft et al. 2018). For this study, we included 185,225 individuals (1,636 PD patients and 183,589 healthy controls) of European ancestry with both genotyping and whole-exome sequencing (WES) data available. Participants were genotyped using the UKB Axiom Array, and imputation was performed with the Haplotype Reference Consortium and UK10K + 1000 Genomes reference panels. Whole-exome sequencing was conducted using the IDT xGen Exome Research Panel v1.0 (Wang et al. 2021). PD diagnosis was based on self-reports by participants or the International Classification of Diseases (ICD-10) diagnosis codes. This included individuals with self-reported code 1262 or ICD-10 code of G20 in hospitalization records. Quality control (QC) followed standard procedures. We excluded outliers with putative sex chromosome aneuploidy (field 22019), high heterozygosity or missing genotype rates (field 22027), and discordant reported versus genotypic sex (field 22001). The analysis was restricted to unrelated individuals to the second degree. The dataset is available for research purposes, and all participants provided documented consent. UKB analyses were conducted using a protocol approved by the Partners HealthCare Institutional Review Board. All study participants provided written informed consents.

### The Luxembourg Parkinson’s Study

For independent replication, we used 653 PD patients and 767 healthy controls from LuxPark (Hipp et al. 2018; Landoulsi et al. 2023), a longitudinal monocentric study within the framework of the NCER-PD (National Centre for Excellence in Research in PD). Genotyping was carried out using the NeuroChip platform (Blauwendraat et al. 2017), while *GBA1* variants were identified using the *GBA1*-targeted PacBio sequencing method (Pachchek et al. 2023). QC for the genotyping data has been previously described (Landoulsi et al. 2023). Participants carrying pathogenic single nucleotide variants (SNVs) or copy number variants (CNVs) in PD-associated genes (*SNCA*, *LRRK2*, *VPS35*, *PRKN*, *PINK1* and *PARK7*) were excluded (Landoulsi et al. 2023). Additionally, we excluded individuals harboring variants of uncertain significance (VUS) or synonymous variants in the *GBA1* gene. All subjects gave written informed consent. The study was approved by the National Research Ethics Committee (CNER Ref: 201407/13).

### Classification of *GBA1* variants

We classified *GBA1* variants based on their pathogenicity in relation to PD and GD, categorizing them as risk, mild, or severe, following the classification of Höglinger and colleagues (Höglinger et al. 2022). Variants identified as pathogenic for GD were classified as either mild or severe for PD, with severe variants exhibiting an odds ratio (OR) of 10–15 for developing PD, while mild variants had an OR of ≤5 (Iwaki et al. 2019; Straniero et al. 2020). Common variants not considered pathogenic for GD but known to increase PD risk (e.g., p.E365K, p.T408M) were categorized as risk variants. Frameshift and nonsense variants in *GBA1* were classified as severe. Variants not classified by Höglinger et al. were further categorized (as severe, mild, or risk) according to the classification of Parlar and colleagues (Parlar et al. 2023) using the online *GBA1* variant browser (https://pdgenetics.shinyapps.io/gba1browser/).

### Polygenic risk score

To generate the PRS, we used the summary statistics from a list of 42 SNPs available from a previous meta-analysis of PD (Chang et al. 2017). Variants within the *GBA1* locus, along with the *LRRK2* p.G2019 pathogenic variants, were excluded for PRS calculation. We calculated PRS using PRsice2 (Choi and O’Reilly 2019), taking into account allele-flipping and using the ‘*–no-regress’* and *’–no-clumping’* options along with the default parameters.

### Statistical analysis

To investigate the association between PRS, *GBA1*_PVs_ carrier status, and PD risk, we performed logistic regression and Cox proportional hazards regression with disease occurrence as the outcome variable. Participants were stratified based on their PRS and *GBA1*_PVs_ carrier status. Initially, individuals were categorized into equal tertiles based on their PRS distribution. Those in the lowest tertile were assigned to the low-PRS group, those in the highest tertile group to the high-PRS group, and those in the middle tertile to the intermediate-PRS group.

For both cohorts, ORs were estimated using a logistic regression model, conditioning on covariates such as sex, age at assessment, and the first four principal components. We applied Cox proportional hazard regression model to evaluate time-to-event data and estimate cumulative incidence risk of PD. Afterwards, we additionally incorporated interactions between *GBA1*_PVs_ status and PRS by introducing an interaction term within the logistic regression model. Finally, we further categorized the *GBA1*_PVs_ carriers’ participants into two groups: (1) mild and severe *GBA1*_PVs_ carriers and (2) risk *GBA1*_PVs_ carriers. This classification allowed to evaluate the association between different levels of *GBA1*_PVs_ severity and the risk of developing PD. The reference category is given by non-carriers with intermediate-PRS. We used R v4.2.2 for all statistical analyses.

## Results

### Demographic characteristics and distribution of *GBA1* variants in UKB and LuxPark

After QC, the final UKB dataset included 185,225 individuals with available WES and genotyping data at the analysis time. Descriptive statistics of the study population after filtering are shown in Table 1. The dataset consisted of 1,636 PD cases, with a mean AAO of 64.6 years, and 183,589 healthy control participants, with a mean age at assessment (AAA) of 64.1 years. In the LuxPark replication cohort, a total of 1,430 individuals were included, comprising 653 PD cases with a mean AAO of 62.4 years and 767 healthy controls with a mean AAA of 59.6 years. The LuxPark cohort exhibited a significantly higher proportion of individuals with a positive family history of PD (FH+) compared to the UKB cohort (chi-squared test *P* < 0.01). In UKB, 57 *GBA1*_PVs_ were identified in 9,019 (4.8%) controls and 145 PD cases (8.8%). The mean of AAO for *GBA1*_PVs_ carriers was 66.2±7.2 years for PD patients and the mean of AAA was 56.6±7.97 for healthy controls. Out of the 57 *GBA1*_PVs_, 40 were classified as severe, 14 as mild, and three as risk variants, with risk variants being the most common (n = 7,990 carriers, Table 2). In the LuxPark cohort, targeted PacBio sequencing of the *GBA1* gene previously revealed that 12.1% (77/637) of PD patients carried *GBA1* variants, with 10.5% (67/637) harboring known pathogenic variants, including severe, mild, and risk variants (Pachchek et al. 2023). For this study, we focused on a subset of individuals who were both genotyped and *GBA1* PacBio-sequenced, identifying 99 carriers of *GBA1*_PVs_. This subset comprised 65 PD cases (9.9%) and 34 controls (4.4%). The mean AAO for PD cases was 61.5 ± 11.7 years, while the mean AAA for healthy controls was 59.4 ± 13.2 years. Of the 12 *GBA1*_PVs_ detected, nine were classified as severe, one as mild, and two as risk variants, with risk variants being the most common (n = 68, Table 2).

**Table 1:**
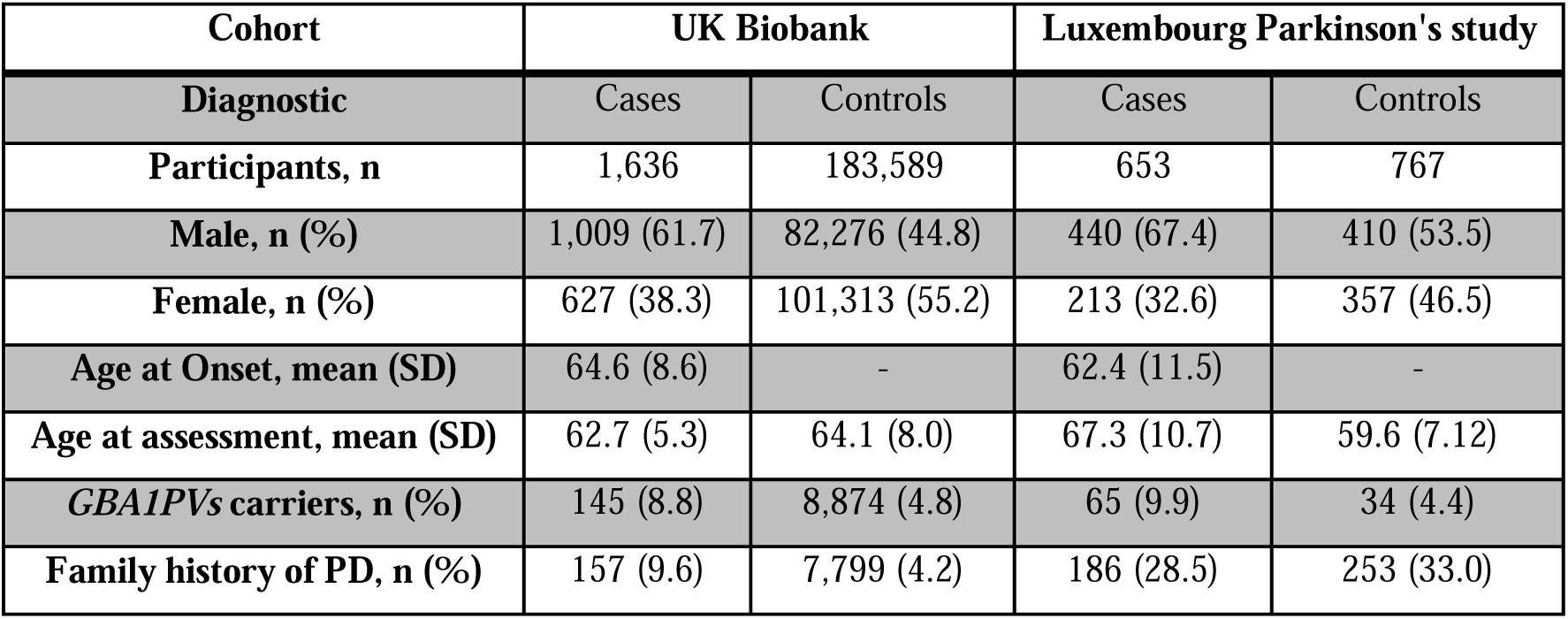
Characteristics of the Luxembourg Parkinson’s Study and the UK Biobank cohort.

| Cohort | UK Biobank |  | Luxembourg Parkinson's study |  |
| --- | --- | --- | --- | --- |
| Diagnostic | Cases | Controls | Cases | Controls |
| Participants, n | 1,636 | 183,589 | 653 | 767 |
| Male, n (%) | 1,009 (61.7) | 82,276 (44.8) | 440 (67.4) | 410 (53.5) |
| Female, n (%) | 627 (38.3) | 101,313 (55.2) | 213 (32.6) | 357 (46.5) |
| Age at Onset, mean (SD) | 64.6 (8.6) | - | 62.4 (11.5) | - |
| Age at assessment, mean (SD) | 62.7 (5.3) | 64.1 (8.0) | 67.3 (10.7) | 59.6 (7.12) |
| *GBA1* PVs carriers, n (%) | 145 (8.8) | 8,874 (4.8) | 65 (9.9) | 34 (4.4) |
| Family history of PD, n (%) | 157 (9.6) | 7,799 (4.2) | 186 (28.5) | 253 (33.0) |

**Table 2:**
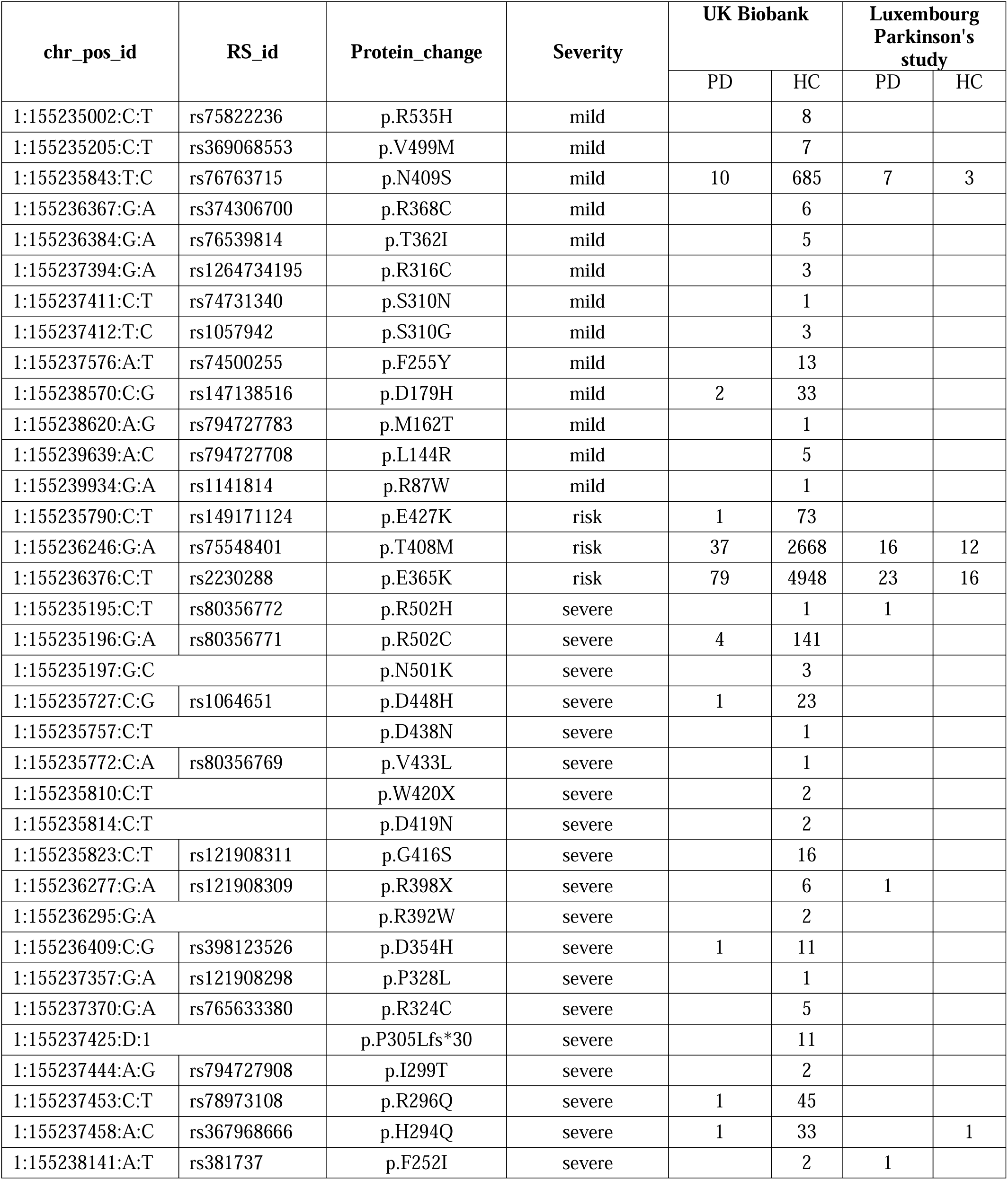
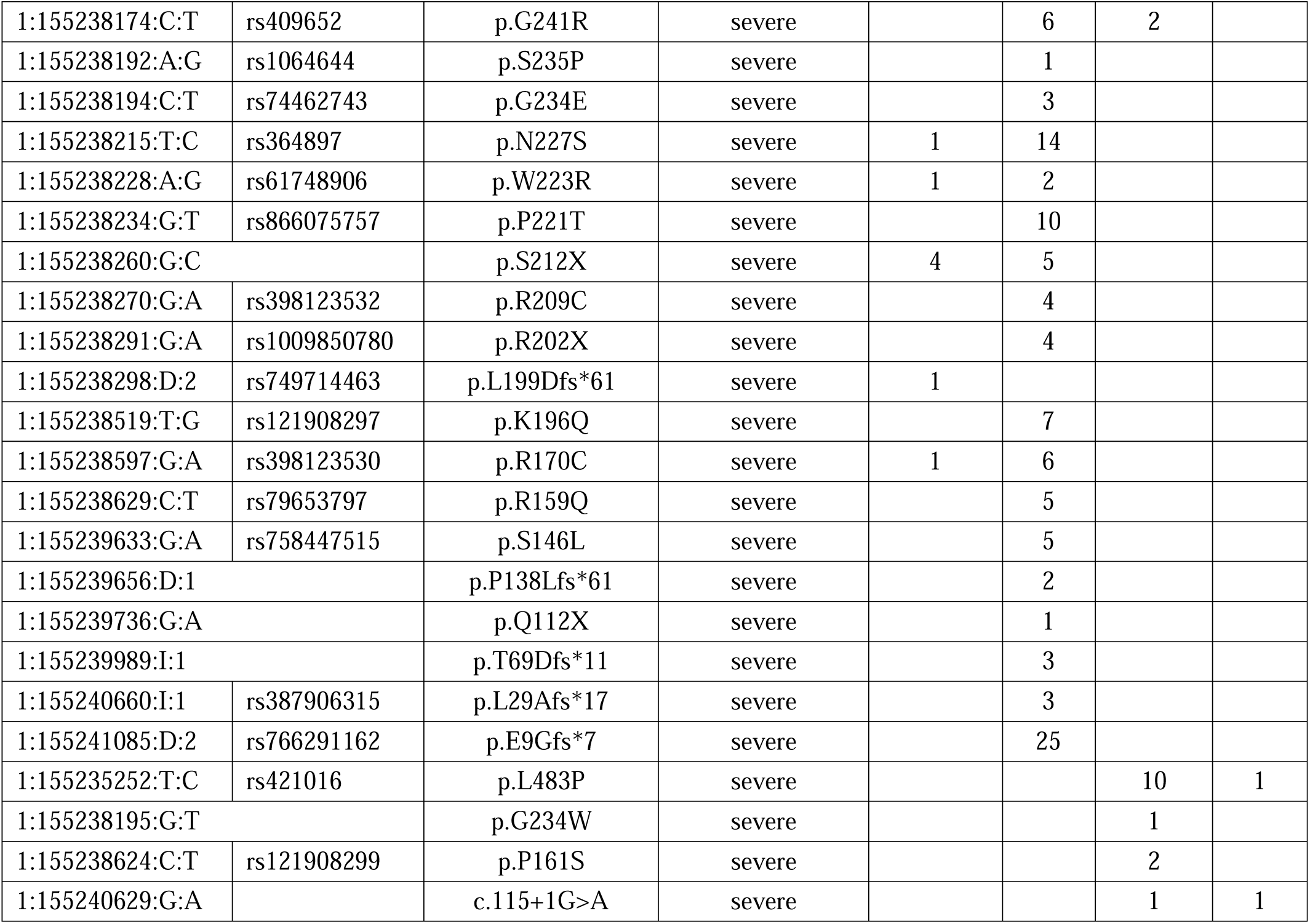
Pathogenic *GBA1* variants used in this study. Abbreviations: PD, Parkinson’s disease; HC, healthy controls.

### Combined effect of PRS and *GBA*_pvs_ status and severity on PD risk

We calculated the PRS using a panel of 42 SNPs to investigate the influence of the genetic background on PD risk in PD *GBA1*_PVs_ carriers. Our analysis revealed that the PRS was significantly higher in PD patients compared to healthy controls in both cohorts (Wilcoxon test *P* < 0.01).

We assessed the influence of PRS and *GBA1*_PVs_ carrier status on PD risk in both cohorts by calculating ORs for PD across PRS categories, using non-carriers with intermediate-PRS as the reference group. PD risk was consistently higher in *GBA1*_PVs_ carriers compared to non-carriers across all PRS categories in both cohorts (Figure 1). In UKB, non-carriers with low- or high-PRS had PD ORs of 0.75 (0.73-0.77) or 1.34 (1.32-1.36) respectively (Figure 1). Among GBA1_PVs_ carriers, those with high-PRS category exhibited OR of 2.34 (95% CI, 2.08-2.63) compared to carriers with low-PRS (OR: 1.13; 95% CI, 0.85-1.49) (Figure 1). Similarly, in LuxPark, *GBA1*_PVs_ carriers with high-PRS had PD ORs of 1.67 (95% CI, 1.55-1.79) compared to those with low PRS (OR: 1.25; 95% CI, 1.07-1.43), although the effect was less pronounced than in UKB. No significant interaction between *GBA1*_PVs_ carrier status and PRS was observed in either UKB (*P* = 0.73) or LuxPark (*P* = 0.48). The Cox proportional hazards analysis confirmed the combined effect of carrier status and PRS on PD risk (Figure 2). Among *GBA1*_PVs_ carriers in UKB, individuals in the high-PRS group showed highest cumulative incidence 67% by the age of 75. In contrast, carriers in the low-PRS group had a lower cumulative incidence, reaching approximately 54% by the same age. At the same age, the cumulative incidence of the disease is consistently higher in *GBA1*_PVs_ carriers compared to non-carriers. Similar trends were observed in LuxPark, the highest cumulative incidence (81%) risk was observed in *GBA1*_PVs_ carriers with high-PRS while ∼75% of individuals within the low-PRS group developed PD at the same age. To gain a deeper understanding of how disease severity is influenced by PRS, we further examined the combined effect of PRS and *GBA1*_PVs_ severity, categorized in two groups: “severe+mild” and “risk”. In both cohorts, we found an association between the severity of *GBA1*_PVs_ and a higher OR for PD (Figure 3). Carriers of severe+mild *GBA1*_PVs_ tended to have higher risk of PD (almost twofold or higher) compared to carriers of risk *GBA1*_PVs_, regardless of their PRS category. In UKB, ORs for carriers of “severe+mild” *GBA1*_PVs_ ranged from 2.05 (95% CI, 1.48-2.83) to 3.69 (95% CI, 2.68-5.04) across the PRS categories, while for carriers of risk *GBA1*_PVs,_ ORs ranged from 1.07 (95% CI, 0.91-1.25) to 2.13 (95% CI, 1.87-2.42, Figure 3). Similarly, in LuxPark, ORs for carriers of “severe+mild” *GBA1*_PVs_ ranged from 1.73 (95% CI, 1.37-2.09) to 1.98 (95% CI, 1.77-2.18) across the PRS categories, while for carriers of risk *GBA1*_PVs,_ ORs ranged from 1.03 (95% CI, 0.79-1.26) to 1.49 (95% CI, 1.29-1.68, Figure 3). No significant interaction between *GBA1*_PVs_ severity (respectively for severe+mild or risk) and PRS was observed in either UKB (*P* = 0.99 and *P* = 0.26) or LuxPark (*P* = 0.44 and *P* = 0.26).

**Figure 1:**
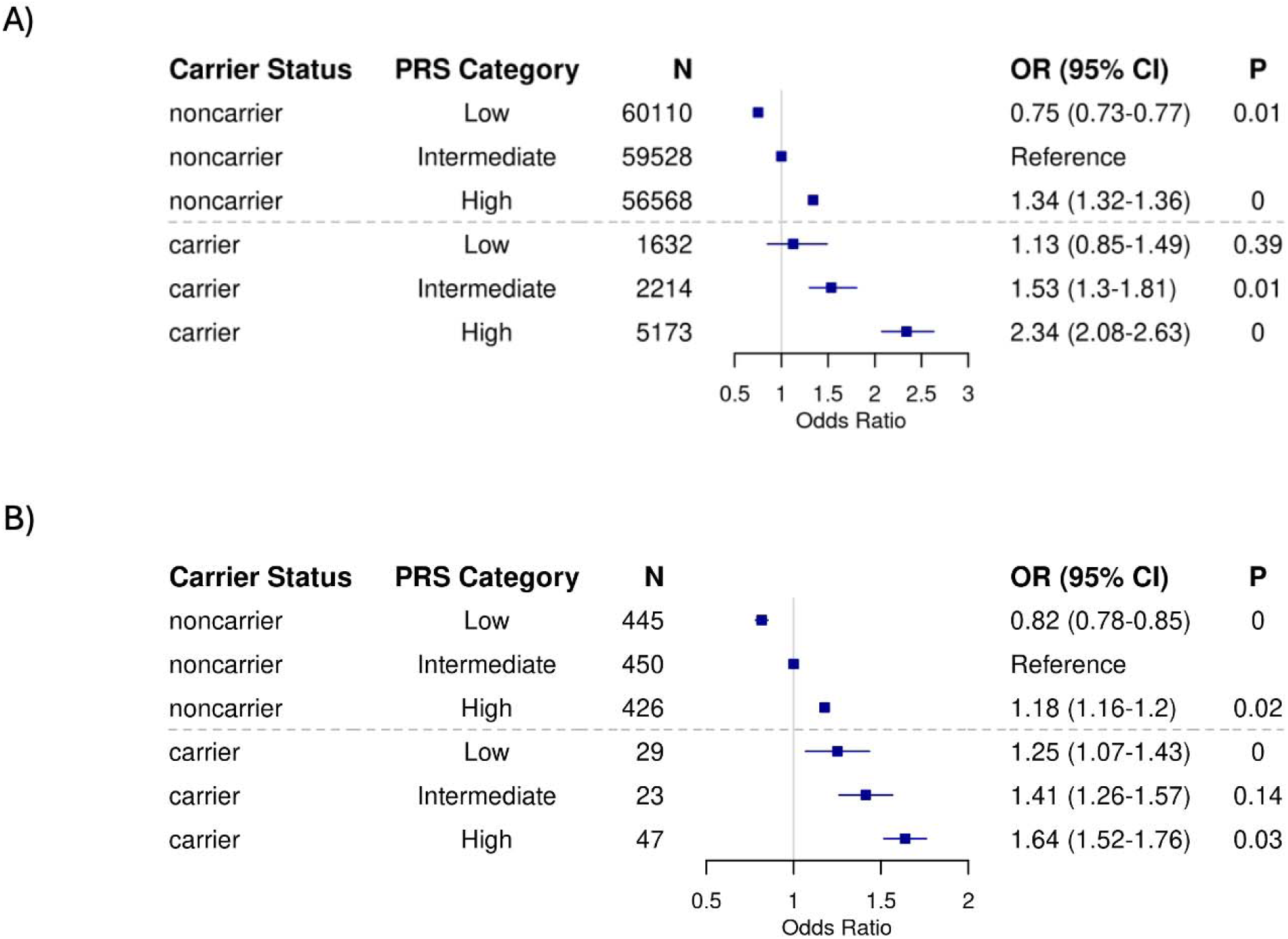
Parkinson’s disease (PD) risk stratified by polygenic risks scores (PRS) categories and *GBA1* carrier status in UK Biobank (A) and the Luxembourg Parkinson’s Study (B) Odds ratio for PD were estimated from logistic models, while conditioning on the sex, age at assessment and the first four ancestry principal components. Non-carriers with median PRS served as the reference group. Carriers and non-carriers were categorized into categories based on their PRS.

**Figure 2:**
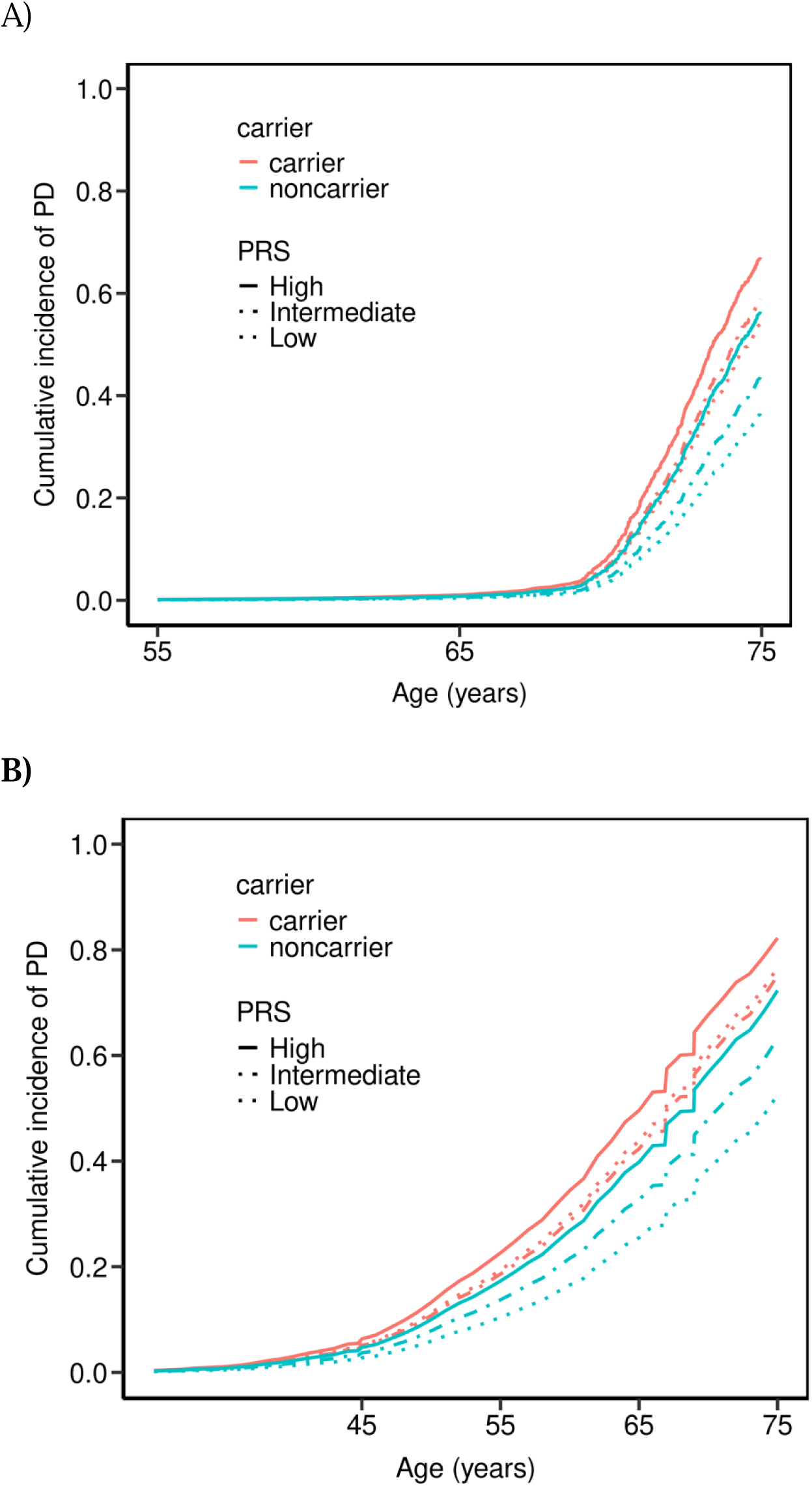
Parkinson’s disease cumulative incidence risk stratified by polygenic risks scores categories and severity status of *GBA1* carrier in UK Biobank (A) and the Luxembourg Parkinson’s Study (B).

**Figure 3:**
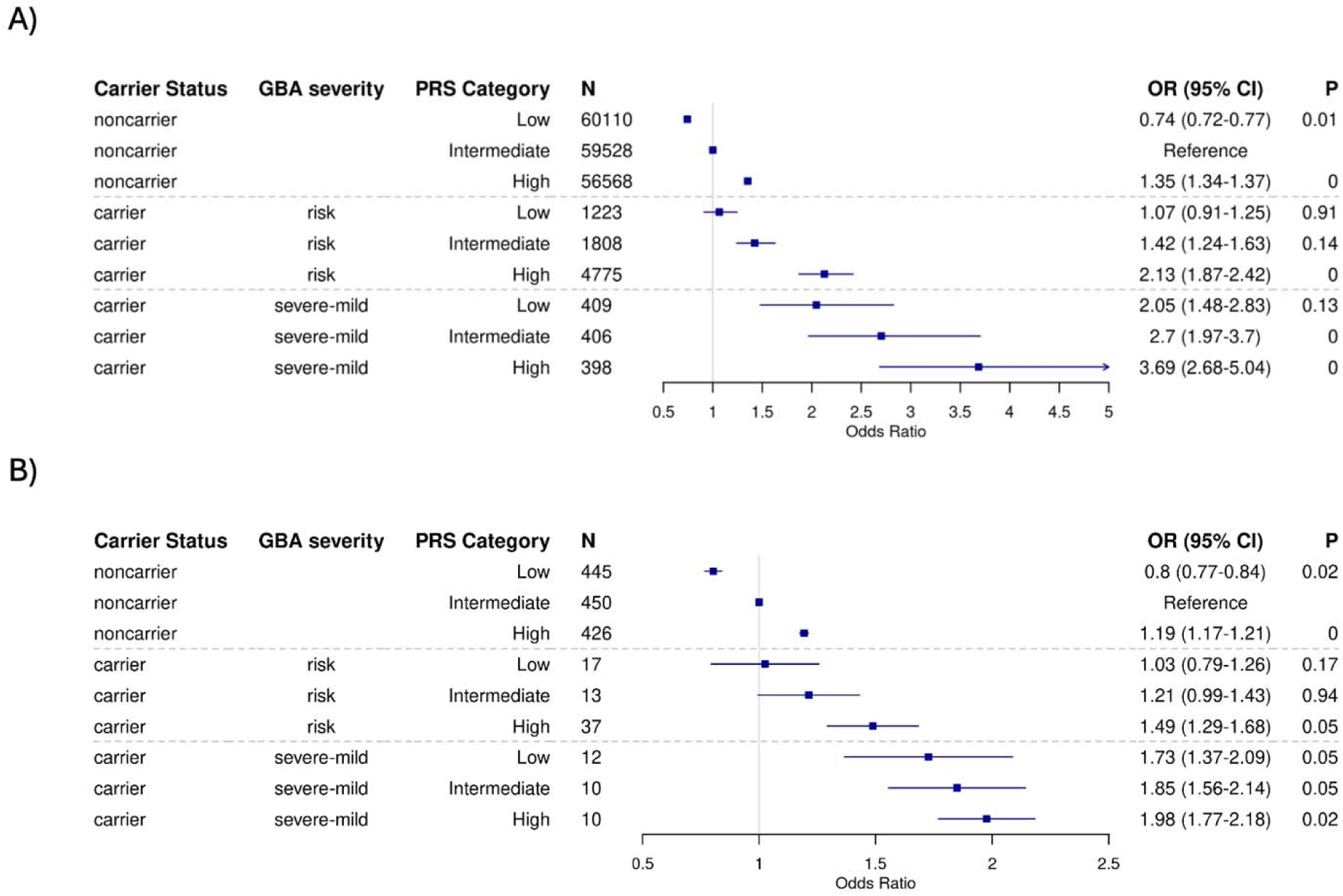
Parkinson’s disease risk stratified by polygenic risks scores categories and severity status of *GBA1* carrier in UK Biobank (A) and the Luxembourg Parkinson’s Study (B).

## Discussion

In this study, we investigated the combined effect of polygenic background and PD-associated *GBA1* variants on the risk of developing PD across two cohorts. Our findings show that both PRSs and *GBA1*_PVs_ independently contribute to PD risk. Individuals carrying *GBA1*_PVs_ consistently showed a higher baseline risk of developing PD compared to non-carriers, regardless of their PRS group. Notably, PRSs independently elevated the odds of developing PD in both carriers and non-carriers. These results suggest that the effects of *GBA1*_PVs_ and PRSs are additive, where the presence of *GBA1*_PVs_ contributes a fixed increase in baseline risk, and PRS independently further increases the overall risk of PD. At older ages, cumulative incidence of PD is consistently higher in *GBA1*_PVs_ compared to non-carriers, therefore, highlighting the increased risk associated with carrying *GBA1*_PVs_. Among *GBA1*_PVs_ carriers, the cumulative incidence is higher in the high-PRS group compared to the low-PRs group, a trend also observed non-carriers. This suggests that PRS exert an additive effect on the penetrance of *GBA1*_PVs_ amplifying the overall risk of PD. The cumulative incidence of PD appears to evolve at an earlier age in the LuxPark cohort compared to the UKB. This difference likely reflects inherent disparities in the study designs and recruitment strategies of the two cohorts. The UKB represents a general population sample, where individuals with early-onset PD are underrepresented. Consequently, the conversion to PD in the UKB cohort may occur later, as the age range and focus of the recruitment potentially exclude younger individuals who are more likely to develop early-onset PD. In contrast, the LuxPark cohort is a PD-specific case-control study, including PD patients with both early-onset and later-onset forms of the disease. As a result, the LuxPark cohort captures a more comprehensive spectrum of PD onset ages, leading to an earlier observed cumulative incidence compared to the UKB.

Using the same approach, we investigated the specific effect of *GBA1*_PVs_ severity, classifying them into two groups: “severe+mild” and “risk” variants. Carriers of severe+mild *GBA1*_PVs_ exhibited the highest baseline risk for developing PD, followed by those with risk variants, further emphasizing the dose-dependent effect of *GBA1*_PVs_ on disease susceptibility. Importantly, PRS continued to elevate PD risk independently and additively, regardless of variants severity.

Our results suggest a clear gradient of risk driven by both *GBA1*_PVs_ severity and PRS. Individuals carrying severe *GBA1*_PVs_ in combination with a high PRS have the highest risk of developing PD, while those with risk variants and a low PRS have a lower, but still elevated, risk compared to non-carriers. Notably, non-carriers with a high PRS also have a substantial risk, but it remains lower than that of any variant carriers with similar PRS levels. These results highlight the complex interplay between polygenic risk and *GBA1*_PVs_, underscoring the importance of considering both factors in genetic assessments of PD risk.

Our findings are consistent with previous studies demonstrating that PRS not only modifies PD risk but also reduces the AAO in carriers of *GBA1* variants, specifically for the two risk variants p.E365K and p.T408M, and the mild p.N409S variant (Blauwendraat et al. 2020). Additionally, variants included in the PD PRS were found to be more frequent in patients with GD type 1 who developed PD, suggesting that common genetic risk variants may influence shared underlying biological pathways (Blauwendraat et al. 2023).

A key finding of this study is that *GBA1* variants and PRS independently and additively increase the risk of PD, with both factors separately contributing to the overall risk. This highlights the importance of considering PRS in clinical trial designs for *GBA1*-related PD, using pre-trial genetic analysis to stratify patients by both *GBA1* variants and overall genetic risk (Blauwendraat et al. 2020; Leonard et al. 2020; Arena et al. 2024). Incorporating PRS alongside whole-exome or targeted gene sequencing in initial diagnoses could streamline healthcare costs, expand cohort sizes, and support the integration of genetic risk data into routine clinical practice (Lewis et al. 2022). Furthermore, combining genetic information with non-genetic factors, such as family history and presymptomatic phenotypes, could improve disease prediction and promote the development of multifactorial risk models (Nalls et al. 2015; Hassanin et al. 2023).

The results of PRS studies can be influenced by factors such as study design and recruitment strategies (Hassanin et al. 2023). In this study, the effects were more pronounced in the discovery cohort (UKB), a population-based sample, compared to the replication cohort (LuxPark), a PD-specific case-control cohort. While both cohorts showed similar trends, the differences are likely due to specific cohort characteristics and recruitment methods. Notably, a significant discrepancy in family history between PD cases and controls was observed in the UKB but not in the Luxembourg cohort, further emphasizing the impact of study design on the outcomes.

This study has several limitations. The UKB cohort is not PD-specific and lacks age and sex matching between cases and controls; however, we mitigated this by including age and sex as covariates in the regression analysis. Additionally, the sample size is limited in certain PRS or *GBA1*_PVs_ severity categories, which may lead to overestimation of effect sizes. These factors should be carefully considered when interpreting the findings.

Overall, this study shows that both PRS and *GBA1*_PVs_ contribute independently and additively to the risk of PD. Carriers of *GBA1*_PVs_ consistently show a higher baseline risk, particularly those with severe variants and high PRS, with PRS influencing *GBA1*_PVs_ penetrance. These results highlight the need to integrate PRS and *GBA1*_PVs_ into PD genetic assessment for better risk stratification. By deepening our understanding of this genetic landscape, especially variant severity, we can pave the way for personalized therapeutic strategies. Ultimately, this approach will bring us closer to tailoring treatments to individual genetic profiles, optimizing outcomes and improving patients’ quality of life.

## Data Availability

All data produced in the present study are available upon request to the respective project.

## Declarations

### Consent for publication

Not applicable.

### Competing interests

No potential conflicts (financial, professional, or personal) relevant to the manuscript.

### Availability of data and materials

Data used to prepare this article were obtained from UKB, and the National Centre of Excellence in Research: Early diagnosis and stratification of Parkinson’s Disease (NCER-PD or the LuxPark https://www.parkinson.lu). Restrictions apply to the availability of these data for UKB, which were used under license for the current study (Project ID: 52446).

### Funding

The FNR supported P.M. as part of the National Centre of Excellence in Research on Parkinson’s disease (NCER-PD, FNR11264123) and the DFG Research Units FOR2715 (INTER/DFG/17/11583046) and FOR2488 (INTER/DFG/19/14429377). The Fonds National de Recherche (FNR) supported D.R.B through the Industrial fellowship program of Luxembourg (FNR14323864).

### Authors’ contributions

(1) Research Project: A. Conception, B. Organization, C. Execution; (2) Statistical Analysis: A. Design, B. Execution; (3) Data: A. acquisition B. Curation (4) Manuscript Preparation: A. Writing of the First Draft, B. Review and Critique.

E.H.: 1A, 1B, 1C, 2A, 2B, 4A, 4B

Z.L.: 3A, 3B, 4A, 4B

S.P.: 3A, 3B, 4A, 4B R.A.: 1A, 4B

R.K.: 3A, 4B

P.K.: 4B

C.M.: 1A, 1B,1C, 4B

P.M.: 1A, 1B, 1C, 3A, 3B, 4A, 4B

D.R.B.: 1A, 1B, 1C, 4A, 4B

## Acknowledgements

Data used to prepare this article were obtained from the UKB, and the National Centre of Excellence in Research: Early diagnosis and stratification of Parkinson’s Disease (NCER-PD) (https://www.parkinson.lu/). Ethics approval for the UK Biobank (UKB) study was obtained from the Northwest Multicentre for Research Ethics Committee (MREC). The UKB ethics statement is available at https://www.ukbiobank.ac.uk/learn-more-about-uk-biobank/about-us/ethics. All UKB participants provided informed consent at recruitment. Similarly, all NCER-PD participants provided written informed consent, and the study was approved by the National Research Ethics Committee (CNER Ref: 201407/13).Parts of the computational analysis were done on the High-Performance Computing cluster of the University of Luxembourg (https://hpc.uni.lu/).

## Supplemental Text

### NCER-PD Consortium Members

Geeta ACHARYA ^2^, Gloria AGUAYO ^2^, Myriam ALEXANDRE ^2^, Muhammad ALI ^1^, Wim AMMERLANN ^2^, Giuseppe ARENA ^1^, Michele BASSIS ^1^, Roxane BATUTU ^3^, Katy BEAUMONT ^2^, Sibylle BÉCHET ^3^, Guy BERCHEM ^3^, Alexandre BISDORFF ^5^, Ibrahim BOUSSAAD ^1^, David BOUVIER ^4^, Lorieza CASTILLO ^2^, Gessica CONTESOTTO ^2^, Nancy DE BREMAEKER ^3^, Brian DEWITT ^2^, Nico DIEDERICH ^3^, Rene DONDELINGER ^5^, Nancy 1. E. RAMIA ^1^, Angelo Ferrari ^2^, Katrin FRAUENKNECHT ^4^, Joëlle FRITZ ^2^, Carlos GAMIO ^2^, Manon GANTENBEIN ^2^, Piotr GAWRON ^1^, Laura Georges ^2^, Soumyabrata GHOSH ^1^, Marijus GIRAITIS ^2,3^, Enrico GLAAB ^1^, Martine GOERGEN ^3^, Elisa GÓMEZ DE LOPE ^1^, Jérôme GRAAS ^2^, Mariella GRAZIANO ^7^, Valentin GROUES ^1^, Anne GRÜNEWALD ^1^, Gaël HAMMOT ^2^, Anne-Marie HANFF ^2,10,11^, Linda HANSEN ^3^, Michael HENEKA ^1^, Estelle HENRY ^2^, Margaux Henry ^2^, Sylvia HERBRINK ^3^, Sascha HERZINGER ^1^, Alexander HUNDT ^2^, Nadine JACOBY ^8^, Sonja JÓNSDÓTTIR ^2,3^, Jochen KLUCKEN ^1,2,3^, Olga KOFANOVA ^2^, Rejko KRÜGER ^1,2,3^, Pauline LAMBERT ^2^, Zied LANDOULSI ^1^, Roseline LENTZ ^6^, Laura LONGHINO ^3^, Ana Festas Lopes ^2^, Victoria LORENTZ ^2^, Tainá M. MARQUES ^2^, Guilherme MARQUES ^2^, Patricia MARTINS CONDE ^1^, Patrick MAY ^1^, Deborah MCINTYRE ^2^, Chouaib MEDIOUNI ^2^, Francoise MEISCH ^1^, Alexia MENDIBIDE ^2^, Myriam MENSTER ^2^, Maura MINELLI ^2^, Michel MITTELBRONN ^1,2,4,10,12,13^, Saïda MTIMET ^2^, Maeva Munsch ^2^, Romain NATI ^3^, Ulf NEHRBASS ^2^, Sarah NICKELS ^1^, Beatrice NICOLAI ^3^, Jean-Paul NICOLAY ^9^, Fozia NOOR ^2^, Clarissa P. C. GOMES ^1^, Sinthuja PACHCHEK ^1^, Claire PAULY ^2,3^, Laure PAULY ^2,10^, Lukas PAVELKA ^2,3^, Magali PERQUIN ^2^, Achilleas PEXARAS ^2^, Armin RAUSCHENBERGER ^1^, Rajesh RAWAL ^1^, Dheeraj REDDY BOBBILI ^1^, Lucie REMARK ^2^, Ilsé Richard ^2^, Olivia ROLAND ^2^, Kirsten ROOMP ^1^, Eduardo ROSALES ^2^, Stefano SAPIENZA ^1^, Venkata SATAGOPAM ^1^, Sabine SCHMITZ ^1^, Reinhard SCHNEIDER ^1^, Jens SCHWAMBORN ^1^, Raquel SEVERINO ^2^, Amir SHARIFY ^2^, Ruxandra SOARE ^1^, Ekaterina SOBOLEVA ^1,3^, Kate SOKOLOWSKA ^2^, Maud Theresine ^2^, Hermann THIEN ^2^, Elodie THIRY ^3^, Rebecca TING JIIN LOO ^1^, Johanna TROUET ^2^, Olena TSURKALENKO ^2^, Michel VAILLANT ^2^, Carlos VEGA ^2^, Liliana VILAS BOAS ^3^, Paul WILMES ^1^, Evi WOLLSCHEID-LENGELING ^1^, Gelani ZELIMKHANOV ^2,3^

^1^ Luxembourg Centre for Systems Biomedicine, University of Luxembourg, Esch-sur-Alzette, Luxembourg.

^2^ Luxembourg Institute of Health, Strassen, Luxembourg.

^3^ Centre Hospitalier de Luxembourg, Strassen, Luxembourg.

^4^ Laboratoire National de Santé, Dudelange, Luxembourg.

^5^ Centre Hospitalier Emile Mayrisch, Esch-sur-Alzette, Luxembourg.

^6^ Parkinson Luxembourg Association, Leudelange, Luxembourg.

^7^ Association of Physiotherapists in Parkinson’s Disease Europe, Esch-sur-Alzette, Luxembourg.

^8^ Private practice, Ettelbruck, Luxembourg.

^9^ Private practice, Luxembourg, Luxembourg.

^10^ Faculty of Science, Technology and Medicine, University of Luxembourg, Esch-sur-Alzette, Luxembourg.

^11^ Department of Epidemiology, CAPHRI School for Public Health and Primary Care, Maastricht University Medical Centre, Maastricht, the Netherlands.

^12^ Luxembourg Center of Neuropathology, Dudelange, Luxembourg.

^13^ Department of Life Sciences and Medicine, University of Luxembourg, Esch-sur-Alzette, Luxembourg.

## Notes

### Competing Interest Statement

The authors have declared no competing interest.

### Author Declarations

Ethics approval for the UK Biobank (UKB) study was obtained from the Northwest Multicentre for Research Ethics Committee (MREC). The UKB ethics statement is available at https://www.ukbiobank.ac.uk/learn-more-about-uk-biobank/about-us/ethics. All UKB participants provided informed consent at recruitment. Similarly, all NCER-PD participants provided written informed consent, and the study was approved by the National Research Ethics Committee (CNER Ref: 201407/13)

## References

Anheim M, Elbaz A, Lesage S, Durr A, Condroyer C, Viallet F, et al. Penetrance of Parkinson disease in glucocerebrosidase gene mutation carriers. Neurology. 2012 Feb 7;78(6):417–20.

Arena G, Landoulsi Z, Grossmann D, Payne T, Vitali A, Delcambre S, et al. Polygenic Risk Scores Validated in Patient Derived Cells Stratify for Mitochondrial Subtypes of Parkinson’s Disease. Annals of Neurology. 2024 Jul;96(1):133–49.

Balestrino R, Tunesi S, Tesei S, Lopiano L, Zecchinelli AL, Goldwurm S. Penetrance of Glucocerebrosidase (*GBA*) Mutations in Parkinson’s Disease: A Kin Cohort Study. Movement Disorders. 2020 Nov;35(11):2111–4.

Blauwendraat C, Faghri F, Pihlstrom L, Geiger JT, Elbaz A, Lesage S, et al. NeuroChip, an updated version of the NeuroX genotyping platform to rapidly screen for variants associated with neurological diseases. Neurobiol Aging. 2017;57:247.e9–247.e13.

Blauwendraat C, Reed X, Krohn L, Heilbron K, Bandres-Ciga S, Tan M, et al. Genetic modifiers of risk and age at onset in GBA associated Parkinson’s disease and Lewy body dementia. Brain. 2020 Jan 1;143(1):234–48.

Blauwendraat C, Tayebi N, Woo EG, Lopez G, Fierro L, Toffoli M, et al. Polygenic Parkinson’s Disease Genetic Risk Score as Risk Modifier of Parkinsonism in Gaucher Disease. Movement Disorders. 2023 May;38(5):899–903.

Bycroft C, Freeman C, Petkova D, Band G, Elliott LT, Sharp K, et al. The UK Biobank resource with deep phenotyping and genomic data. Nature. 2018 Oct;562(7726):203–9.

Chang D, Nalls MA, Hallgrímsdóttir IB, International Parkinson’s Disease Genomics Consortium, 23andMe Research Team, Hunkapiller J, et al. A meta-analysis of genome-wide association studies identifies 17 new Parkinson’s disease risk loci. Nat Genet. 2017 Oct;49(10):1511–6.

Choi SW, O’Reilly PF. PRSice-2: Polygenic Risk Score software for biobank-scale data. GigaScience. 2019 Jul 1;8(7):giz082.

Gan-Or Z, Liong C, Alcalay RN. GBA-Associated Parkinson’s Disease and Other Synucleinopathies. Curr Neurol Neurosci Rep. 2018 Aug;18(8):44.

Hassanin E, Spier I, Bobbili DR, Aldisi R, Klinkhammer H, David F, et al. Clinically relevant combined effect of polygenic background, rare pathogenic germline variants, and family history on colorectal cancer incidence. BMC Med Genomics. 2023 Mar 5;16(1):42.

Hipp G, Vaillant M, Diederich NJ, Roomp K, Satagopam VP, Banda P, et al. The Luxembourg Parkinson’s Study: A Comprehensive Approach for Stratification and Early Diagnosis. Front Aging Neurosci. 2018 Oct 29;10:326.

Höglinger G, Schulte C, Jost WH, Storch A, Woitalla D, Krüger R, et al. GBA-associated PD: chances and obstacles for targeted treatment strategies. J Neural Transm. 2022 Sep;129(9):1219–33.

Iwaki H, Blauwendraat C, Leonard HL, Liu G, Maple-Grødem J, Corvol JC, et al. Genetic risk of Parkinson disease and progression: An analysis of 13 longitudinal cohorts. Neurol Genet. 2019 Aug;5(4):e348.

Landoulsi Z, Pachchek S, Bobbili DR, Pavelka L, May P, Krüger R, et al. Genetic landscape of Parkinson’s disease and related diseases in Luxembourg. Front Aging Neurosci. 2023 Dec 20;15:1282174.

Leonard H, Blauwendraat C, Krohn L, Faghri F, Iwaki H, Ferguson G, et al. Genetic variability and potential effects on clinical trial outcomes: perspectives in Parkinson’s disease. J Med Genet. 2020 May;57(5):331–8.

Lewis ACF, Perez EF, Prince AER, Flaxman HR, Gomez L, Brockman DG, et al. Patient and provider perspectives on polygenic risk scores: implications for clinical reporting and utilization. Genome Med. 2022 Oct 7;14(1):114.

Nalls MA, Blauwendraat C, Vallerga CL, Heilbron K, Bandres-Ciga S, Chang D, et al. Identification of novel risk loci, causal insights, and heritable risk for Parkinson’s disease: a meta-analysis of genome-wide association studies. The Lancet Neurology. 2019 Dec;18(12):1091–102.

Nalls MA, McLean CY, Rick J, Eberly S, Hutten SJ, Gwinn K, et al. Diagnosis of Parkinson’s disease on the basis of clinical and genetic classification: a population-based modelling study. The Lancet Neurology. 2015 Oct;14(10):1002–9.

Pachchek S, Landoulsi Z, Pavelka L, Schulte C, Buena-Atienza E, Gross C, et al. Accurate long-read sequencing identified GBA1 as major risk factor in the Luxembourgish Parkinson’s study. npj Parkinsons Dis. 2023 Nov 23;9(1):156.

Parlar SC, Grenn FP, Kim JJ, Baluwendraat C, Gan Or Z. Classification of *GBA1* Variants in Parkinson’s Disease: The *GBA1* PD Browser. Movement Disorders. 2023 Mar;38(3):489–95.

Rizig M, Bandres-Ciga S, Makarious MB, Ojo OO, Crea PW, Abiodun OV, et al. Identification of genetic risk loci and causal insights associated with Parkinson’s disease in African and African admixed populations: a genome-wide association study. The Lancet Neurology. 2023 Nov;22(11):1015–25.

Sidransky E, Nalls MA, Aasly JO, Aharon-Peretz J, Annesi G, Barbosa ER, et al. Multicenter analysis of glucocerebrosidase mutations in Parkinson’s disease. N Engl J Med. 2009 Oct 22;361(17):1651–61.

Straniero L, Asselta R, Bonvegna S, Rimoldi V, Melistaccio G, Soldà G, et al. The SPID-GBA study: Sex distribution, Penetrance, Incidence, and Dementia in GBA-PD. Neurol Genet. 2020 Dec;6(6):e523.

Wang Q, Dhindsa RS, Carss K, Harper AR, Nag A, Tachmazidou I, et al. Rare variant contribution to human disease in 281,104 UK Biobank exomes. Nature. 2021 Sep 23;597(7877):527–32.

